# Perceived stress by students of the medical sciences in Cuba toward the COVID-19 pandemic: results of an online survey

**DOI:** 10.1101/2020.12.16.20248345

**Authors:** Frank Hernández-García, Onelis Góngora Gómez, Victor Ernesto González-Velázquez, Elys María Pedraza-Rodríguez, Rolando Zamora-Fung, Luis Alberto Lazo Herrera

## Abstract

**Introduction:** The aims of this study were to determine the usefulness of the Modified (10-items) Scale of Perceived Stress related to COVID-19 (EEP-10-C by its acronym in Spanish) and to identify the levels of stress perceived by students of the medical sciences in Cuba toward the COVID-19 pandemic.

**Methods:** A cross-sectional study was conducted, with self-reported data of students from fourteen Cuban Universities of Medical Sciences (n = 200), through an online survey. The EEP-10-C was used as an instrument to identify stress. Its validity was determined through a confirmatory factor analysis and its internal consistency and reliability was measured by the Cronbach’ alph. A cluster analysis was performed to establish as cut-off point the center of the cluster with the highest values of stress perceived by the scale.

**Results:** The average age of the sample was 23.30 ± 1.91 years, with observed scores of the EEP-10-C between 0 and 29 points (13.25 ± 5,404). When applying the cut-off point ≥25, only two students had high rates of stress perceived. The confirmatory factor analysis supported the validity of the instrument; with a Cronbach’s alpha of 0.755. The cutoff point ≥20 was proposed as a reference of high stress perceived for the study population, when applying this one, 14% of students presented high rates of stress.

**Discussion:** In Cuba, students of the medical sciences have participated in research and supporting health care, despite which they have presented low levels of stress. The main contribution of the research was the validation of the EEP-10-C for its use in assessing levels of stress in Cuban medical students, proposing the cut-off point ≥20 as a reference of high stress perceived.

## Introduction

The appearance of the Severe Acute Respiratory Syndrome Coronavirus 2 (SARS-CoV-2) that causes the coronavirus disease (COVID-19), declared as a pandemic by the World Health Organization (WHO) on March 11, 2020 [1], has resulted in a succession of changes in all spheres of social life. We must acknowledge that the senior educational system was also unprepared for a disruption of the kind of the COVID-19 pandemic [2]; consequently more than 100 countries have been affected by the suspension of teaching activities [1].

Medical education has been affected as well in different regions of the planet [3, 4] and the proposals made to mitigate the effects of the discontinuity of teaching are numerous, reaching from the implementation of distance education modalities, to the incorporation of students to health care labors [5-10]; in such a way that students becomes an active element in solving the problem and not part of it.

In Cuba, students of finals years of the medical careers have been worked bringing assistance in hospitals and isolation centers for cases suspicious of COVID-19. In addition, some distance teaching modalities have been trialed with good results so far, regarding that they have been never used in Cuban medical education. An element to be emphasized is the massive incorporation of students from all medical careers and academic years to active inquiries work in primary health care, in search of respiratory symptomatic cases, with prior vigorous training for this activity [11-13].

Stress is a widespread health problem in the world [13]. It is the process or reflex that is set in motion when a person perceives a complex situation in which they find something that is threatening themselves. According to its determination, it is a social phenomenon, and by its nature, it is also a psychophysiological singularity [15], which by acting constantly on one individual and added to other factors, becomes a trigger for numerous diseases. University students constitute a vulnerable population for mental health problems because of the changes they experiment in the transition to adult life [16,17].

The COVID-19 pandemic has generated among the population high degrees of stress, anxiety and depression, resultant from the effects of quarantine and social isolation, to which it is added for the specific case of health professionals, the stress generated by health care to the contagious patients and overload of work [18-20]. Regarding these elements, it is necessary to have valid and reliable measurements implemented in a remote manner. In this sense, among other instruments, there is the Cohen scale of stress perceived (PSE) [21] and a modified version of it for the Colombian population [22]. The last one, known as the 10-item stress perceived scale modified in relation to COVID-19 (EEP-10-C), more adapted to the Latin American context, have not been validated among students of the medical sciences or other populations than the Colombian.

In the actual Cuban context, there is an absence of a validated instrument to determine the stress levels perceived toward the COVID-19 pandemic, and specifically among students of the medical sciences, who as well as the health professionals, are individuals with high risk of infeccion. It worth it to investigate if the scale based on the EEP-10-C is valid among students of the medical sciences, regarding the characteristics of this kind of population, as well as the possibility of facilitating more specific evaluations and interventions in the vulnerable population. The aims of this study were to asses the usefulness of the EPP-10-C scale and to identify the stress levels perceived in Cuban students of the medical sciences during the COVID-19 pandemic.

## Methods

### Design and study population

An observational, analytical and cross-sectional, multicenter study was carried out. Data was self-reported data and collected through social networks. The group of possible participants was made up of students of the medical sciences (medicine, stomatology, nursing and health technologies) of the 2019-2020 academic year, from 14 of the 16 Medical Universities of Cuba. Of all the potential participants, 200 students completed and returned the surveys.

### Data Processing

The questionnaires were sent through groups of medical science students on WhatsApp. Before sending the survey, the participants gave their consent by reading and approving the study objectives, the target population and the nature of their voluntary participation, the risks/benefits and the confidentiality of their data. The survey was sent on August 4, 2020 at 20:00 hrs and the responses were expected until August 17 at 20:00 hours.

### Instrument and measurements

Data were collected on the following sociodemographic traits of the participants: age, gender, academic year and Medical University. In order to determine possible stressors associated with stress due to the COVID-19 pandemic, the following information was requested: cohabitation at home, medical history, history of relative infected by COVID-19. Finally, the labor carried out during the months of the pandemic/quarantine was collected (“permanently isolated at home”, “related to investigation work”, “providing health care in hospitals”, “collaborating in an isolation centers for suspected patients”, “other activity” and specify, or a combination of the above).

Then, in the same survey, participants answered the version modified by Campo-Arias [22] of the 10-item stress perceived scale modified in relation to COVID-19. This scale was previously adjusted in its items 2 and 6 and with language adapted to our context. The EEP-10-C is composed of 10 items; each one of them offers 5 response options: “never”, “almost never”, “occasionally”, “almost always” and “always”. Items 1, 3, 9 and 10 are scored directly from 0 to 4 and items 2, 4, 5, 6, 7 and 8, contrariwise, from 4 to 0. The EEP-10-C was considered due to the acceptable internal consistency shown in other studies [21]. The suggested high stress cut-off point perceived by the EEP-10-C developers for the population of the scale creators was ≥25 points.

### Analysis of data

The Monte Carlo significance for Pearson’s Chi Square test was determined to determine whether there were statistical differences between genderes in responses to the multiple choice survey. The validity of the instrument was determined by an exploratory factor analysis (EFA) using the Kaiser-Meyer-Olkin (KMO) sampling adequacy measure and Bartlett’s sphericity test to identify whether the items grouped a latent factor. The internal consistency and reliability of the scale was calculated with Cronbach’s alpha coefficient. A K-means cluster analysis was conducted, regarding the total score of the EPP-10-C scale as the dependent variable, to establish as the cut-off point the center of the cluster with the highest values of stress perceived by the scale. The means between groups were compared using the Mann-Whitney U test after verifying the non-normality of the distribution using the Kolmogorov-Smirnov test. Data analysis was performed using IBM-SPSS version 22.0.

### Ethical principles

Research was carried out in accordance with the Declaration of Helsinki. There was no potential harm to participants, informed consent forms were collected and anonymity was guaranteed.

## Results

**Table 1** shows the descriptive analysis of the sample, composed of students from all Medical Universities of Cuba. There was representativeness of all careers, with a wild predominance of Medicine. Students in terminal years (fourth, fifth and sixth) constituted the majority of the group studied, with predominance of young students who stioll lived with their parents. In the medical history analysis, it was found that bronchial asthma was reported by 28% of the students, while 60% did not report the presence of personal pathological antecedents. None of the students referred to have been ill with COVID-19, however 7.5% of those surveyed reported knowing someone who contracted the disease. The active inquiries work predominated among the activities carried out during the epidemic months (77.0%), where only 1.5% of the students reported having been permanently isolated at home.

**Table 1.**
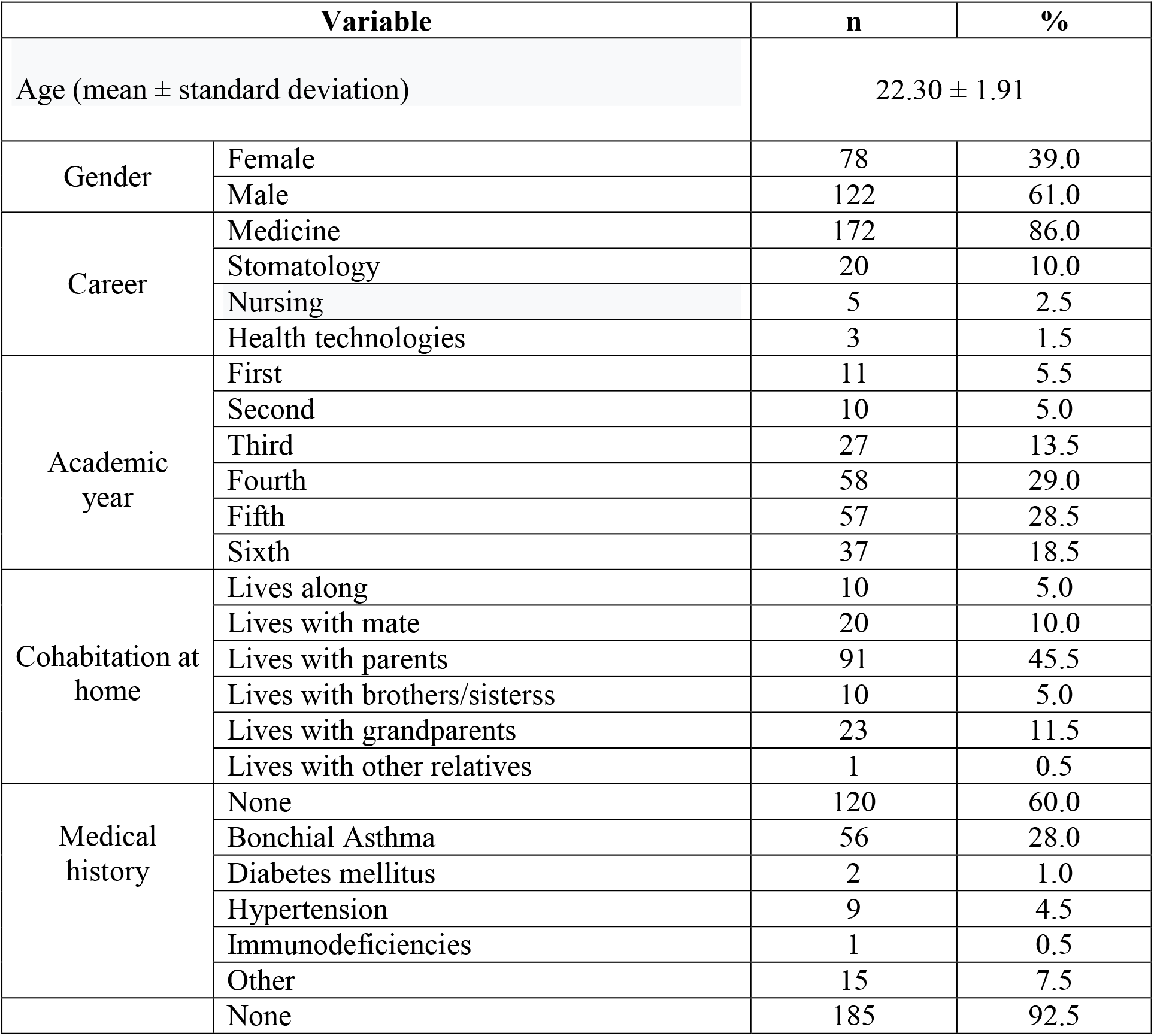
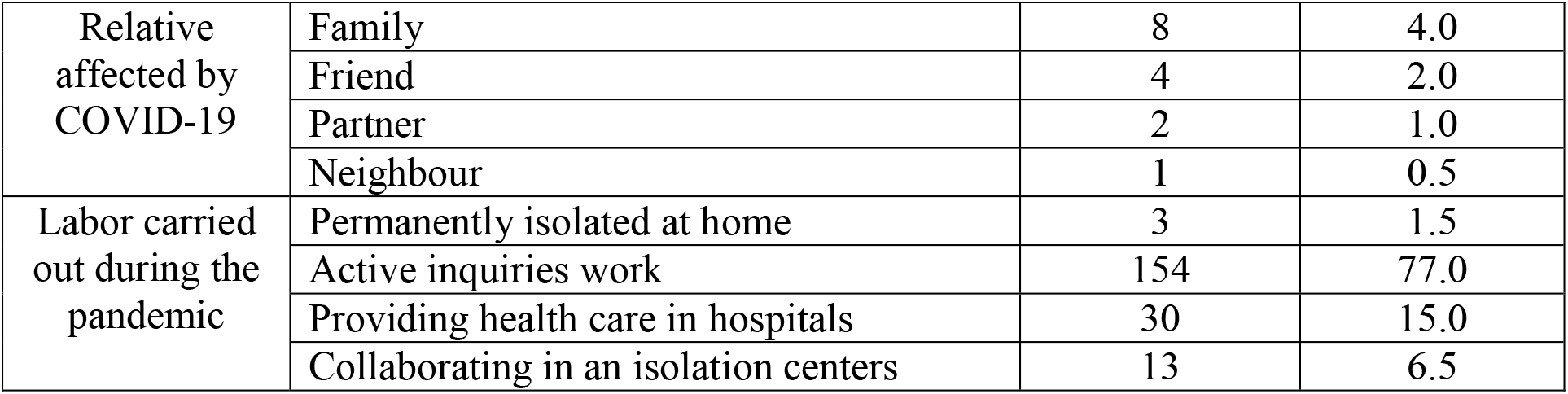
Socio-educational characteristics of students of the medical sciences who participated in the online survey.

**Table 2** shows that there were no statistically significant differences between genders in terms of answers given in the EEP-10-C questionnaire in any of the items that are evaluated on the scale. The scores obtained on the scale were generally low. Only two subjects scored higher than the cut-off point (25) proposed previously for high stress perceived related to COVID-19. The mean of the scale scores was 13.25 ± 5,404, with values between 0 and 29 points.

**Table 2.** Distribution of the score on the 10-item stress perceived scale modified in relation to COVID-19 (EEP-10-C) regarding gender.

| During the last month | Never |  | Hardly ever |  | Occasionally |  | Almost always |  | Always |  | *p*<sup>*</sup> |
| --- | --- | --- | --- | --- | --- | --- | --- | --- | --- | --- | --- |
|  | M<br>n (%) | F<br>n (%) | M<br>n (%) | F<br>n (%) | M<br>n (%) | F<br>n (%) | M<br>n (%) | F<br>n (%) | M<br>n (%) | F<br>n (%) |  |
| 1. I have been affected as if something serious will happen unexpectedly with the epidemic. | 38<br>(31.1) | 21<br>(26.9<br>) | 33<br>(27.0) | 15<br>(19.2<br>) | 45<br>(36.9) | 36<br>(46.2) | 4<br>(3.3) | 6 (7.7) | 2 (1.6) | 0<br>(0.0) | 0.22<br>4 |
| 2. I have felt able to control the important things in my life because of the epidemic. | 1 (0.8) | 1<br>(1.3) | 9 (7.4) | 7<br>(9.0) | 21<br>(17.2) | 13<br>(16.7) | 57<br>(46.7<br>) | 32<br>(41.0) | 34<br>(27.9) | 25<br>(32.1<br>) | 0.92<br>9 |
| 3. I have felt nervous or stressed about the epidemic | 11<br>(9.0) | 4<br>(5.1) | 29<br>(23.8) | 13<br>(16.7<br>) | 51<br>(41.8) | 33<br>(42.3) | 20<br>(16.4<br>) | 20<br>(25.6) | 11<br>(9.0) | 8<br>(10.3<br>) | 0.38<br>0 |
| 4. I have been confident about my ability to handle my personal problems related to the epidemic | 2 (1.6) | 0 (0.0) | 3 (1.5) | 1 (1.3) | 22 (18.0) | 11 (14.1) | 42 (34.4 ) | 32 (41.0) | 53 (43.4) | 34 (43.6 ) | 0.65<br>9 |
| 5. I have felt that things are going well (optimistic) with the epidemic | 3 (2.5) | 5 (6.4) | 13 (10.7) | 11 (14.1 ) | 50 (41.0) | 37 (47.4) | 38 (31.1 ) | 20 (25.6) | 18 (14.8) | 5 (6.4) | 0.19<br>2 |
| 6. I have felt able to face the things that I have to do to control the possible infection | 1 (0.8) | 0 (0.0) | 3 (2.5) | 2 (2.6) | 10 (8.2) | 12 (15.4) | 53 (43.4 ) | 27 (34.6) | 55 (45.1) | 37 (47.4 ) | 0.43<br>7 |
| 7. I have felt that I can control the difficulties that could appear in my life due to the infection | 1 (0.8) | 3 (3.8) | 6 (4.9) | 4 (5.1) | 29 (23.8) | 16 (20.5) | 54 (44.3 ) | 33 (42.3) | 32 (26.2) | 22 (28.2 ) | 0.66<br>9 |
| 8. I have felt that I have almost everything under control regarding the epidemic | 3 (2.5) | 7 (9.0) | 10 (8.2) | 8 (10.3 ) | 40 (32.8) | 27 (34.6) | 51 (24.8 ) | 26 (33.3) | 18 (14.8) | 10 (12.8 ) | 0.25<br>7 |
| 9. I have been upset that things related to the epidemic are almost all out of control | 15 (12.3) | 10 (12.8 ) | 18 (14.8) | 11 (14.1 ) | 58 (47.5) | 28 (35.9) | 26 (21.3 ) | 19 (24.4) | 5 (4.1) | 10 (12.8 ) | 0,16<br>1 |
| 10. I have felt that difficulties accumulate in these days of the epidemic and I feel unable to overcome them | 41 (33.6) | 30 (38.5 ) | 42 (34.4) | 20 (25.6 ) | 31 (25.4) | 18 (23.1) | 5 (4.1) | 7 (9.0) | 3 (2.5) | 3 (3.8) | 0.44<br>1 |

F: female, M: male, \* Bilateral asymptotic significance of the Monte Carlo test.

The measurement instrument validity was confirmed by the exploratory factor analysis (EFA), which made it possible to determine that the sample is adequate for the instrument, with association between items. In this analysis, it was found that all the communalities in the extraction were greater than 0.4; with a sample adequacy measure of KMO > 0.5 and a statistical significance of Bartlett sphericity test < 0.05 **(Table 3)**. These results allow to applie the scale legitimately regarding its validation in the analyzed sample.

**Table 3.** Exploratory factor analysis according to the Kaiser-Meyer-Olkin sample adequacy measure.

|  |  |  |
| --- | --- | --- |
| Kaiser-Meyer-Olkin sampling adequacy measure |  | 0.770 |
| Bartlett's sphericity | Estimated Chi Square | 389.437 |
|  | df | 45 |
|  | *p* | 0.000 |
| **Extraction method:** Main components analysis **df:** degrees of freedom. |  |  |

**Table 4** shows the reliability analysis by applying Cronbach’s alpha coefficient, which allows to evaluate the internal consistency of the instrument. The internal consistency of the EEP-10-C is good (Cronbach’s Alpha = 0.755), which means that the scale has good psychometric properties, representing the high rate to which the items are correlated with each other. The results of the descriptive reliability analysis for Cronbach’s Alpha if one element is eliminated, did not show any useful results under the assumption of eliminating any of the items to increase the reliability of the scale, thus confirming the validity and precision of the EEP-10-C.

**Table 4.** Reliability analysis of the 10-item stress perceived scale modified in relation to COVID-19 (EPP-10-C).

| Items | Mean of scale if item is removed | Scale variance if item is removed | Corrected item-total correlation | Cronbach's alpha if item is removed |
| --- | --- | --- | --- | --- |
| 1. I have been affected as if something serious will happen unexpectedly with the epidemic. | 12.01 | 24.332 | 0.412 | 0.735 |
| 2. I have felt able to control the important things in my life because of the epidemic. | 12.19 | 23.951 | 0.478 | 0.726 |
| 3. I have felt nervous or stressed about the epidemic | 11.22 | 24.213 | 0.378 | 0.740 |
| 4. I have been confident about my ability to handle my personal problems related to the epidemic | 12.45 | 24.178 | 0.509 | 0.723 |
| 5. I have felt that things are going well (optimistic) with the epidemic | 11.57 | 24.106 | 0.439 | 0.731 |
| 6. I have felt able to face the things that I have to do to control the possible infection | 12.54 | 25.335 | 0.402 | 0.737 |
| 7. I have felt that I can control the difficulties that could appear in my life due to the infection | 12.14 | 22.982 | 0.600 | 0.709 |
| 8. I have felt that I have almost everything under control regarding the epidemic | 11.73 | 23.959 | 0.429 | 0.733 |
| 9. I have been upset that things related to the epidemic are almost all out of control | 11.27 | 24.922 | 0.287 | 0.755 |
| 10. I have felt that difficulties accumulate in these days of the epidemic and I feel unable to overcome them | 12.15 | 25.023 | 0.292 | 0.753 |
| **Cronbach's alpha = 0.755** |  |  |  |  |

**Table 5** shows the clusters obtained through the K-means analysis, by means of which three homogeneous but at the same time significantly different from each other groups were created (p = 0.000) according to the score on the EEP-10-C scale. The centers of the final clusters represent the average values of each cluster, therefore it is understood as the mean score obtained by the subjects belonging to each group.

**Table 5.** Cluster analysis of K-means according to the score on the scale of stress perceived related to the COVID-19 pandemic (EPP-10-C).

| EEP-10-C scores | Centers of the finals conglomerates |  |  | Individuals in each conglomerated |  |  | Square mean | df | ANOVA |  |
| --- | --- | --- | --- | --- | --- | --- | --- | --- | --- | --- |
|  | 1 | 2 | 3 | 1 | 2 | 3 |  |  | F | p |
|  | 6 | 13 | 20 | 52 | 92 | 56 | 2435.37 | 2 | 509.98 | 0.000* |
| *Statistically significant. **df:** degrees of freedom. |  |  |  |  |  |  |  |  |  |  |

Cluster 3 was made up of the 56 individuals with the highest scores on the EEP-10-C scale, so its center is proposed as a reference to establish the cut-off point (≥ 20) for high stress perceived related to COVID-19 in the study population.

Table 6 shows the comparison of the sample according to the score achieved on the scale and the classification of the groups using the proposed cut-off point (≥20). It was observed that 28 students were identified with high stress perceived related to COVID-19, resulting in the difference between both groups as statistically significant (p = 0.000).

**Table 6.** Distribution of students according to scores on the 10-item stress perceived scale modified in relation to COVID-19 (EPP-10-C) and stress levels according to the cut-off point ≥ 20.

|  | High stress<br>perceived | N | Meab | Standard<br>deviation | Tipic error<br>of mean | *p* |
| --- | --- | --- | --- | --- | --- | --- |
| **EPP-10-C scores** | No | 172 | 11.88 | 4.458 | 0.340 | 0.000* |
|  | Yes | 28 | 21.68 | 1.926 | 0.364 |  |
| *Statistically significant. |  |  |  |  |  |  |

## Discussion

This is the first study applying the 10-item stress perceived scale modified in relation to COVID-19 (EPP-10-C) [22] or some version of the Cohen scale [21] in medical science students in Cuba and in students from Spanish America in general. Participants in this investigation were all young students of the medical sciences, the majority of whom were male. However, no statistically significant differences were found between gender and responses to the EEP-10-C or high level of stress, which differs from other studies [23, 24].

In this investigation, when applying the cut-off point ≥25, only two students had a high degree of stress perceived, with a mean score in the EEP-10-C of 13.25 ± 5.40 points, which indicated low stress perceived in this population. Campo-Arias [22] obtained a mean score on the scale of 16.5 ± 7.3, with values between 0 and 36 points, reporting 58 (14.28%) individuals with high stress perceived, but it should be noted that its population was more heterogeneous regarding age, with participants between 19 and 88 years old (43.9 ± 12.4). Aslan and Pekince [24] evaluated stress perceived in nursing students in Turkey, using a validated version of Cohen’s scale, in which they obtained a mean of score on the scale much higher than the present investigation (31.69 ± 6.91).

Other authors have evaluated the psychological impact of COVID-19. In a study that evaluated the stress perceived in students at KSA using the Cohen scale [25], mean scores of 22.12 ± 7.33 were shown, which are higher compared to this study, and concerned a higher degree of stress. Similar results to those have been found in researchs carried out on university students in France [26], Spain [27], Belarus and Russia [28] during the COVID-19 pandemic, where anxiety, stress and depression were reported. The results of these studies may be partly due to the stress generated in the context of quarantine and social distancing [19], which has affected the whole world, but more severely to countries with higher population density.

Students of the medical sciences in Cuba have been actively participating in labors of antagonizing the disease related to active inquiries work or providing health care. On the other hand, this fact generates a paradox, because although they are not victims of the severe effects of prolonged quarantine and social distancing, they have been exposed to a major risk, given by the low percentage of stress perceived (1% for cut-off point ≥25 and 14% for cut-off point ≥20). This last result may be influenced by having up-to-date information on the disease, which has been associated with less psychological impact [18, 29].

Not all studies has shown a negative impact of COVID-19 on the mental health of university students. An investigation carried out in China on undergraduate medical students showed that only a minority of this group had moderate (2.7%) or severe (0.9%) anxiety [30].

After performing the K-means conglomerate analysis, from which a cut-off point of ≥20 is proposed in the EEP-10-C for Cuban medical science students, those classified as high stress perceived increase, despite this new group only represent 14% of the total. In another study carried out in Cuba, it was found that medical students who participated in the active inquiries work presented low vulnerability to stress, not being present in 83% of the sample studied [31]. This result is similar to the obteined in this study, though in that case the instruments used was different from ours and it was not validated for the population on the context of COVID-19.

The limitations of the study include those inherent to cross-sectional studies when establishing statistical associations and not causality. Other limitations include the procedures for the conformation of the sample (non-probabilistic) and the virtual form of the survey that could generate recall and selection biases, as well as the short time that it was arranged to be answered by the students. In contrast, stress perceived was not stratified into levels, only the high degree of stress perceived or not was considered. Future research should take this limitation into account and stratify the population according to the level of stress into low, medium and high levels and look for possible statistical associations between this and sociodemographic factors, which was not possible in this research due to the poor percentage of stress perceived (<1%) and even after adjusting the cut-off point to ≥ 20 (14%), however, the validity and internal consistency of the EEP-10-C was demonstrated, with the potentiality to generalize its application in Cuba.

The strength of the research is that it is one of the first studies to address aspects of the mental and occupational health of students of the medical sciences in Cuba toward the epidemic, carried out with an easy-to-fill online survey. Another strength is the representativeness of students from all the Medical Universities of Cuba. The main contribution of the research was the validation of the EEP-10-C for Cuban medical majors and also the proposal of the cut-off point ≥20 as high stress perceived for this population to be evaluated for other authors in future research.

Other subsequent studies should evaluate the validity of the instrument for the rest of the Cuban population and health professionals, to assess if it is necessary to formulate psychological interventions to improve the mental health of vulnerable populations during the COVID-19 epidemic.

## Data Availability

the authors are in possession of all data.

## Acknowledgments

Our most sincere thanks to students of the medical sciences in Cuba who voluntarily agreed to participate in this survey.

## Conflicts of interest

The authors declare no conflicts of interest.

## Notes

### Competing Interest Statement

The authors have declared no competing interest.

### Funding Statement

No one

### Author Declarations

Ethics Comitee of Universidad de Ciencias Medicas de Ciego de Avila

## References

1. Alemán I, Vera E, Patiño-Torres MJ. COVID-19 y la educación médica: retos y oportunidades en Venezuela. Educ Med. 2020; 21(4): 272–276.

2. Pedró F. COVID-19 y educación superior en América Latina y el Caribe: efectos, impactos y recomendaciones políticas. Análisis Carolina. 2020; (36): 1–15.

3. Choi B, Jegatheeswaran L, Minocha A, Alhilani M, Nakhoul M, Mutengesa E. The impact of the COVID-19 pandemic on final year medical students in the United Kingdom: a national survey. BMC Med Educ. 2020; 20: 206.

4. Klasen JM, Vithyapathy A, Zante B, Burm S. “The storm has arrived”: the impact of SARS-CoV-2 on medical students. Perspect Med Educ. 2020; 9: 181–185.

5. Valdez-García JE, López Cabrera MV, Jiménez Martínez MA, Díaz Elizondo JA, Gerardo Dávila Rivas JA, Olivares Olivares SL. Me preparo para ayudar: respuesta de escuelas de medicina y ciencias de la salud ante COVID-19. Inv Ed Med. 2020; 9(35): 85–95.

6. San-Juan-Bosch M, García-Núñez R, Mur-Villar N, Falcón-Hernández A, Díaz-Brito A. Experiencias y alternativas académicas de la Universidad de Ciencias Médicas de Cienfuegos durante la COVID-19. Medisur. 2020; 18(3): 410–415.

7. Artopoulos A. COVID-19: ¿Qué hicieron los países para continuar con la educación a distancia?. Revista Latinoamericana de Educación Comparada. 2020; 11(17): 1–11.

8. Vergara de la Rosa E, Vergara Tam R, Alvarez Vargas M, Camacho Saavedra L, Galvez Olortegui J. Educación médica a distancia en tiempos de COVID-19. Educ Méd Super. 2020; 34(2): e2383.

9. Vitón-Castillo AA, Lazo Herrera LA. Las TIC en la educación médica cubana en tiempos de COVID-19. Educ Med. 2020.

10. Tabari P, Amini M, Moosavi M. Lessons learned from COVID-19 epidemic in Iran: The role of medical education. Med Teach. 2020; 42(7): 833.

11. Hernández-García F, Góngora-Gómez O. Rol del estudiante de ciencias médicas frente a la COVID-19: el ejemplo de Cuba. Educ Med. 2020.

12. Falcón-Hernández A, Navarro-Machado V, Díaz-Brito A, Delgado-Acosta H, Valdés-Gómez M. Pesquisa activa masiva poblacional para la COVID-19. Experiencia con estudiantes de las ciencias médicas. Cienfuegos, 2020. Medisur. 2020; 18(3): 381–387.

13. Pedraza-Rodríguez EM, González-Velázquez VE, López-Baeza PM. Labor de los estudiantes de medicina cubanos en la pesquisa activa durante la pandemia del SARS-CoV-2. Unimed. 2020; 2(3): 1–3.

14. Alfonso Águila B, Calcines Castillo M, Monteagudo de la Guardia R, Nieves Achon Z. Estrés académico. Rev EDUMECENTRO. 2015; 7(2): 163–178.

15. Pérez Núñez D, García Viamontes J, García González TE, Ortiz Vázquez D, Centelles Cabreras M. Conocimientos sobre estrés, salud y creencias de control para la Atención Primaria de Salud. Rev Cubana Med Gen Integr. 2014; 30(3): 354–363.

16. Auerbach RP, Mortier P, Bruffaerts R, Alonso J, Benjet C, Cuijpers P, et al. WHO World mental health surveys international college student project: prevalence and distribution of mental disorders. J Abnorm Psychol. 2018; 127(7): 623–638.

17. Micin S, Bagladi V. Salud Mental en Estudiantes Universitarios: Incidencia de Psicopatología y Antecedentes de Conducta Suicida en Población que Acude a un Servicio de Salud Estudiantil. Ter Psicol. 2011; 29(1): 53–64.

18. Lozano-Vargas A. Impacto de la epidemia del Coronavirus (COVID-19) en la salud mental del personal de salud y en la población general de China. Rev Neuropsiquiatr. 2020; 83(1): 51–56.

19. Brooks SK, Webster RK, Smith LE, Woodland L, Wessely S, Greenberg N. The psychological impact of quarantine and how to reduce it: rapid review of the evidence. Lancet. 2020; 395: 912–920.

20. Qiu J, Shen B, Zhao M, Wang Z, Xie B, Xu Y. A nationwide survey of psychological distress among Chinese people in the COVID-19 epidemic: implications and policy recommendations. General Psychiatry. 2020; 2(33): 20200306.

21. Cohen S, Kamarck T, Mermelstein R. A global measure of stress perceived. J Health Soc Behav. 1983; 24(4): 385–396.

22. Campo-Arias A, Pedrozo-Cortés MJ, Pedrozo-Pupo JH. Escala de estrés percibido relacionado con la pandemia de COVID-19: Una exploración del desempeño psicométrico en línea. Rev Colomb Psiquiatr. 2020.

23. Matus E, Matus L, Florez AM, Stanziola M, Araguás N, López A, et al. Estrés por COVID-19 en Panamá. Alternativas cubanas en Psicología. 2020; 8(24): 120–135.

24. Aslan H, Pekince H. Nursing students’ views on the COVID-19 pandemic and their percieved stress levels. Perspect Psychiatr Care. 2020.

25. AlAteeq DA, Sumayah Aljhani S, AlEesa D. Stress perceived among students in virtual classrooms during the COVID-19 outbreak in KSA. J Taibah Univ Med Sci. 2020.

26. Husky MM, Kovess-Masfety V, Swendsen JD. Stress and anxiety among university students in France during Covid-19 mandatory confinement. Compr Psychiatry. 2020; 102: 152191.

27. Odriozola González P, Planchuelo Gómez A, Irurtia MJ, de Luis García R. Psychological effects of the COVID 19 outbreak and lockdown among students and workers of a Spanish University. Psychiatry Res. 2020; 290: 113108.

28. Gritsenko V, Skugarevsky O, Konstantinov V, Khamenka N, Marinova T, Reznik A, et al. COVID 19 Fear, Stress, Anxiety, and Substance Use Among Russian and Belarusian University Students. Int J Ment Health Addiction. 2020.

29. Wang C, Pan R, Wan X, et al. Immediate Psychological Responses and Associated Factors during the Initial Stage of the 2019 Coronavirus Disease (COVID-19) Epidemic among the General Population in China. Int J Environ Res Public Health. 2020; 17(5): 1729.

30. Cao W, Fang Z, Hou G, Han M, Xu X, Dong J, et al. The psychological impact of the COVID-19 epidemic on college students in China. Psychiatry Res. 2020; 287: 112934.

31. Pérez Abreu MR, Gómez Tejeda JJ, Tamayo Velázquez O, Iparraguirre Tamayo AE, Besteiro Arjona ED. Alteraciones psicológicas en estudiantes de medicina durante la pesquisa activa de la COVID-19. MEDISAN. 2020; 24(4): 537–548.

